# Surgical Risk Assessment and Outcomes in Transthyretin Amyloidosis Cardiomyopathy

**DOI:** 10.64898/2026.07.10.26357789

**Authors:** Karan Shahi, Shawn Sud, Robert JH. Miller, James A. White, Nowell M. Fine

**Affiliations:** Division of Cardiology, Department of Cardiac Sciences, Libin Cardiovascular Institute, University of Calgary, Calgary, Alberta, Canada; Division of General Internal Medicine, Department of Medicine, University of Calgary, Calgary, Alberta, Canada

**Author notes:** Address for Correspondence: Nowell Fine, South Health Campus Hospital, 4448 Front Street SE Calgary, Alberta, Canada, T3M 1M4.

**Keywords:** postoperative, risk stratification, transthyretin amyloidosis cardiomyopathy

## Abstract

**Background:** Transthyretin amyloidosis cardiomyopathy (ATTR-CM) is an infiltrative cardiomyopathy and an increasingly recognized cause of heart failure. With improved survival from disease-modifying therapies, an increasing number of patients are presenting for surgery and may be at increased risk of adverse postoperative outcomes. This study reports outcomes of ATTR-CM patients undergoing surgery and evaluates the utility of the Revised Cardiac Risk Index (RCRI), a perioperative risk tool.

**Methods:** A total of 145 ATTR-CM patients were included, among which 51 patients underwent at least one eligible surgical procedure. Preoperative risk was assessed using the RCRI, analyzed both as a categorical and as a dichotomized (≥3 vs <3) variable. Postoperative outcomes included unplanned hospital admission, length of stay (LOS), prolonged hospitalization (>48 hours), and major adverse cardiac events. Models were adjusted for frailty (Clinical Frailty Scale ≥5) and major surgery, using multivariable, ordinal, and Firth penalized logistic regression analyses.

**Results:** Patients were predominantly male (86%) with a mean age of 76 ± 9 years, and 61% were frail. Higher RCRI scores were associated with unplanned postoperative hospital admission (RCRI ≥3: adjusted OR 48.9, 95% CI 4.8–502.2) and longer LOS (RCRI ≥3: adjusted OR 40.7, 95% CI 4.3–382.8). RCRI ≥3 was also associated with prolonged hospitalization (>48 hours) in Firth penalized logistic regression, whereas frailty was not independently associated.

**Conclusions:** In a real-world ATTR-CM cohort undergoing major non-cardiac surgery, the overall risk of adverse outcomes was low, and higher RCRI scores were associated with increased postoperative hospital admission and longer LOS, including hospitalization exceeding 48 hours. The RCRI retains prognostic utility in this high-risk cohort and may support peri-operative risk stratification.

## INTRODUCTION

Transthyretin amyloidosis cardiomyopathy (ATTR-CM) is an infiltrative cardiomyopathy caused by progressive deposition of misfolded transthyretin (TTR) proteins within the myocardial interstitium.^1,2^ ATTR-CM has two disease subtypes: hereditary (hATTR), resulting from pathogenic TTR gene mutations, and wild-type (wtATTR), which occurs in the absence of a mutation and predominantly affects older individuals.^1,2^ ATTR-CM is an increasingly recognized cause of heart failure in the aging population.^1^

Beyond heart failure, ATTR-CM is associated with atrial arrhythmias, conduction system disease, aortic stenosis, and autonomic dysfunction.^1,3^ These features may increase peri-operative vulnerability in patients undergoing cardiac and non-cardiac surgery.^4^ Furthermore, individuals with ATTR-CM are frequently older and have a substantial burden of comorbidities, potentially compounding surgical risk. With the availability of disease-modifying therapies, improving disease awareness and increasing diagnosis rates, a growing number of patients with ATTR-CM may present for cardiac and non-cardiac surgical procedures.^4,5^ However, peri-operative risk stratification in this population remains poorly defined. Existing surgical risk models are derived from general populations, and their applicability to ATTR-CM has not been established.

The Revised Cardiac Risk Index (RCRI) is a widely used, guideline-endorsed tool for estimating peri-operative cardiovascular risk before planned surgery.^6,7^ In broader surgical cohorts, higher RCRI scores are associated with increased postoperative complications and longer hospitalization.^6^ Whether the RCRI retains predictive utility in patients with ATTR-CM is unknown. It is also unclear whether the RCRI can identify ATTR-CM patients at risk of prolonged hospitalization beyond standard postoperative monitoring thresholds, commonly defined as 48 hours in higher-risk cardiac patients (RCRI ≥3).^7,8^

The purpose of this study was to describe postoperative outcomes and evaluate the performance of the RCRI in patients undergoing cardiac and non-cardiac surgery, with assessment of its association with; (1) postoperative hospital admission, (2) length of stay (LOS), including prolonged hospitalization exceeding 48 hours, and (3) occurrence of major adverse cardiovascular events (MACE). We hypothesized that higher RCRI scores would be associated with greater postoperative monitoring needs and longer hospital stay in this high-risk population.

## METHODS

A total of 145 patients with a diagnosis of ATTR-CM followed at the Cardiac Amyloidosis Clinic at the University of Calgary (Calgary, Alberta, Canada) between May 2011 and November 2021 were initially identified. Of these, 51 patients underwent at least one relevant surgical procedure (major non-cardiac surgery, major cardiac surgery, or minor non-cardiac procedure) and formed the study cohort for peri-operative analysis. ATTR-CM diagnosis was established according to consensus guidelines, either by biopsy tissue with proteomic analysis using mass spectrometry or by technetium-99m-pyrophosphate (Tc-99m-PYP) nuclear bone scintigraphy with single-photon emission computed tomography (SPECT), after exclusion of light-chain (AL) amyloidosis.^9^ Genetic testing was performed in all patients to differentiate ATTR-CM subtypes. The study was approved by the University of Calgary Research Ethics Board with a waiver of the requirement for patient informed consent.

### Data Element Collection

#### Clinical Data Collection

Baseline clinical, laboratory, and imaging data were extracted from the Cardiovascular Imaging Registry of Calgary (CIROC, NCT04367220) a comprehensive cardiovascular clinical outcomes registry (Libin Cardiovascular Institute, University of Calgary).^10^ The date of ATTR-CM diagnosis, defined as the date of positive Tc-99m-PYP imaging or confirmatory biopsy, was used to extract baseline data. Missing or additional data not available in CIROC were manually abstracted from patient medical records.

#### Surgical Procedures

All surgical procedures performed after ATTR-CM diagnosis were identified through manual abstraction of patient medical records. All surgical procedures requiring intraoperative anesthesiologist care and monitoring, including those performed under general and regional/neuraxial anesthesia were included.

Minor non-cardiac procedures requiring sedation common to the ATTR-CM population, such as carpal tunnel release were excluded from analyses. Cardiac procedures requiring sedation such as pacemaker and implantable cardioverter defibrillator (ICD) implantation, right heart catheterizations, and endomyocardial biopsies, were also excluded. Procedures not expected to impose significant cardiovascular or systemic stress (e.g., otolaryngologic, ophthalmological, or endoscopic procedures) were not included. Emergency surgeries/procedures were excluded. All cases with uncertainty regarding eligibility were reviewed by the study team (S.S., K.S., and N.M.F).

#### Surgical Parameters

Pre-operative risk was assessed using RCRI for all major non-cardiac surgeries.^6^ RCRI was analyzed primarily as a categorical variable (0–6, with 6 representing the highest risk level) and secondarily as a dichotomous variable (≥3 vs <3).^6,11^ For cardiac surgical procedures, pre-operative risk was descriptively assessed using the Society of Thoracic Surgeons (STS) risk score, including estimated operative mortality and morbidity/mortality risk.^12^

#### Postoperative Data and Outcomes

Postoperative outcomes collected included MACE within 30 days, intensive care unit (ICU) admission and duration, hospital length of stay, postoperative complications, invasive interventions/reoperations, and 30-day readmissions. MACE included non-fatal myocardial infarction, non-fatal stroke, or mortality. Cause of death was adjudicated as cardiovascular or non-cardiovascular using ICD-10 coding and medical record review (N.M.F). Postoperative complications were graded according to the Clavien-Dindo classification system (I–V), which categorizes complications based on the severity of intervention required.^13,14^ Postoperative disposition outcomes included three measures. Postoperative admission was defined as same-day discharge versus unplanned hospital stay of ≥24 hours. Length of stay (LOS) category was classified as same-day discharge, 24–48 hours, or >48 hours. The primary outcome, prolonged hospitalization, was defined as unplanned LOS >48 hours, surpassing the recommended postoperative monitoring threshold. This threshold aligns with peri-operative guideline-recommended postoperative surveillance periods (approximately 48-72 hours) for high-risk patients undergoing non-cardiac surgery, during which routine monitoring for peri-operative myocardial injury and cardiac events is typically performed.^7,8^

### Statistical Analyses

Continuous variables that were normally distributed are presented as mean ± standard deviation (SD), while non-normally distributed variables are reported as median with interquartile range (IQR). For normally distributed data, two-sample or paired t-tests were used as appropriate. For non-normally distributed data, the Mann-Whitney U test was used for independent groups, and the Wilcoxon signed-rank test for paired groups. Categorical variables were expressed as absolute frequencies and relative percentages, and comparisons were performed using Fisher’s exact test. A two-sided p-value of less than 0.05 was considered statistically significant.

As there were only n=2 cardiac surgical procedures identified, these were reported descriptively and were excluded from inferential analyses due to insufficient sample size for modelling. Accordingly, all regression analyses were restricted to major non-cardiac surgical procedures.

Associations between RCRI and postoperative outcomes were explored using univariable methods: standard logistic regression for postoperative admission (same-day discharge vs ≥24 hours) and ordinal logistic regression for LOS category (same-day discharge, 24–48 hours, or >48 hours). Clinically relevant covariates, including frailty (clinical frailty scale; CFS ≥5)^15^ and major non-cardiac surgery, were included in all multivariable models. The primary outcome, prolonged hospitalization (LOS >48 hours), was uncommon, creating a risk of separation with standard logistic regression. To address this, Firth penalized logistic regression was used to reduce small-sample bias and provide stable estimates.^16,17^

RCRI was analyzed primarily as a categorical variable (1, 2, ≥3) to preserve the clinical risk gradient, with secondary analyses using a dichotomized RCRI (≥3 vs <3) for sensitivity analyses. Final multivariable models adjusted for frailty and major non-cardiac surgery. Effect estimates from all multivariable models are reported as odds ratios (ORs) with 95% confidence intervals (CIs). All analyses were performed using STATA Version 17, Basic Edition (College Station, TX, USA).

## RESULTS

### Patient Characteristics

A total of 145 patients diagnosed with ATTR-CM were initially identified. Of these, 51 patients (44 male, 86%; mean age at diagnosis 76 ± 9 years) underwent at least one eligible surgical procedure (major non-cardiac surgery, major cardiac surgery, or minor non-cardiac procedure) and formed the final study cohort for peri-operative analyses (**Table 1**). Patients who did not undergo any surgical procedure or who underwent only excluded minor cardiac procedures (as described above) were not included in the surgical cohort. Overall, 37 patients (73%) were on disease-modifying therapy. Six patients (12%) had hATTR, and the remainder had wtATTR. At baseline, 16 patients (31%) were classified as New York Heart Association (NYHA) functional class III–IV. Atrial fibrillation or flutter was present in 36 patients (71%), and 13 (25%) had a pacemaker or implantable cardioverter-defibrillator (ICD). Frailty was prevalent, with 31 patients (61%) classified as frail (CFS ≥5).

**Table 1.**
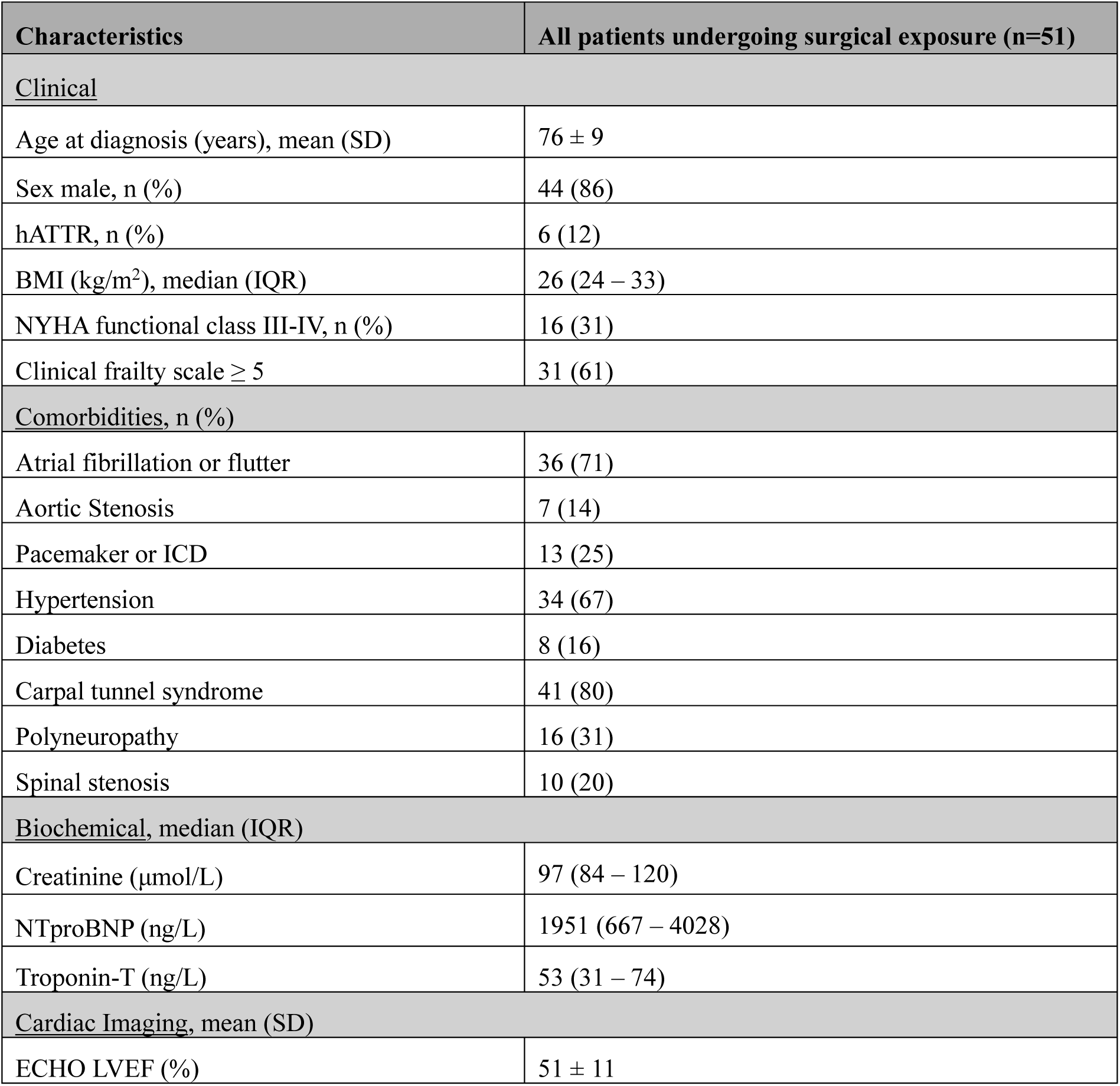
Baseline transthyretin amyloidosis cardiomyopathy (ATTR-CM) patient characteristics among patients undergoing surgical exposure. BMI-body mass index, ECHO-transthoracic echocardiography, hATTR-hereditary transthyretin amyloidosis, ICD-implantable cardioverter-defibrillator, IQR-interquartile range, LVEF-left ventricular ejection fraction, NTproBNP-N-terminal pro B-type natriuretic peptide, NYHA-New York Heart Association, SD-standard deviation.

| Characteristics | All patients undergoing surgical exposure (n=51) |
| --- | --- |
| <u>Clinical</u> |  |
| Age at diagnosis (years), mean (SD) | 76 ± 9 |
| Sex male, n (%) | 44 (86) |
| hATTR, n (%) | 6 (12) |
| BMI (kg/m<sup>2</sup>), median (IQR) | 26 (24 – 33) |
| NYHA functional class III-IV, n (%) | 16 (31) |
| Clinical frailty scale ≥ 5 | 31 (61) |
| <u>Comorbidities, n (%)</u> |  |
| Atrial fibrillation or flutter | 36 (71) |
| Aortic Stenosis | 7 (14) |
| Pacemaker or ICD | 13 (25) |
| Hypertension | 34 (67) |
| Diabetes | 8 (16) |
| Carpal tunnel syndrome | 41 (80) |
| Polyneuropathy | 16 (31) |
| Spinal stenosis | 10 (20) |
| <u>Biochemical, median (IQR)</u> |  |
| Creatinine (μmol/L) | 97 (84 – 120) |
| NTproBNP (ng/L) | 1951 (667 – 4028) |
| Troponin-T (ng/L) | 53 (31 – 74) |
| <u>Cardiac Imaging, mean (SD)</u> |  |
| ECHO LVEF (%) | 51 ± 11 |

### Surgical Characteristics

A total of 72 surgical procedures were performed in this cohort (51 patients). Surgical procedure categories are described in **Table 2**. The number of procedures per patient was: 1 (n=35), 2 (n=12), 3 (n=3), and 4 (n=1). Among the 72 procedures, 48 were discharged on the same day, and 24 (33%) required at least a 24-hour stay (**Table 3**). The average length of stay among admitted patients was 5.5 ± 6.5 days.

**Table 2.** Surgical exposure among patients with transthyretin amyloidosis cardiomyopathy (ATTR-CM). Surgical exposures were not mutually exclusive; patients may appear in more than one category.

| Variable | n (%) |
| --- | --- |
| ≥1 cardiac surgery | 2 (3.9) |
| ≥1 major non-cardiac surgery | 37 (72.5) |
| ≥1 minor non-cardiac surgery | 17 (33.3) |

**Table 3.** Operative characteristics and 30-day postoperative outcomes. MACE-major adverse cardiovascular event. SD-standard deviation.

| Variable | n (%) |
| --- | --- |
| <u>Type of Surgery</u> |  |
| Cardiac | 2 (2.8) |
| Major non-cardiac | 51 (70.8) |
| Minor non-cardiac | 19 (26.4) |
| <u>Anesthesia Type</u> |  |
| General | 30 (41.7) |
| Regional/neuraxial | 19 (26.4) |
| Monitored sedation | 5 (6.9) |
| Local/other | 18 (25.0) |
| <u>Postoperative Hospitalization & Outcomes</u> |  |
| Same-day discharge | 48 (66.7) |
| ≥1-day stay | 24 (33.3) |
| Length of stay among admitted, mean ± SD (days) | 5.5 ± 6.5 |
| Postoperative complications | 8 (11.1) |
| MACE (composite) | 3 (4.2) |

MACE occurred in three surgical procedures (4.2%): two non-fatal myocardial infarctions (one following left total hip arthroplasty and the other following transurethral resection of the prostate), and one non-cardiac death (following right hemicolectomy). ICU admissions and hospital readmissions occurred in one patient each. There were no postoperative invasive interventions or reoperations. Eight procedures were followed by postoperative complications, including seven Clavien–Dindo grade II complications requiring pharmacologic treatment and one grade V complication resulting in death (as described above).

### Cardiac Surgeries

Two patients underwent cardiac surgery. One underwent triple vessel coronary artery bypass grafting and had a 5-day postoperative hospital stay; the estimated Society of Thoracic Surgeons (STS) risk score operative mortality and morbidity/mortality risks were 3.05% and 11.30%, respectively. The second underwent excision of severe mitral annular calcification with reconstruction of the mitral annulus, mitral valve replacement, and septal myectomy, with a postoperative hospital stay of 16-days and estimated operative mortality and morbidity/mortality risks of 11.50% and 28.60%, respectively. Neither patient experienced postoperative MACE.

### Major Non-Cardiac Surgeries

Of the 72 procedures, 51 (71%) were classified as major non-cardiac surgeries. The distribution of RCRI scores was: 1 (n=17), 2 (n=22), and ≥3 (n=12). Seventeen major non-cardiac procedures (33%) were performed in patients meeting CFS criteria (≥5) as frail.

Higher RCRI scores were associated with postoperative hospital admission (≥24 hours) (**Figure 1**). In multivariable logistic regression adjusting for frailty and major non-cardiac procedures, both RCRI score 2 and ≥3 demonstrated substantially increased odds of admission compared with RCRI score 1 (RCRI score 2 adjusted OR 13.0, 95% CI 1.5–113.9, p = 0.02; RCRI ≥3 adjusted OR 48.9, 95% CI 4.8–502.2, p = 0.001) (**Table 4**).

**Figure 1.**
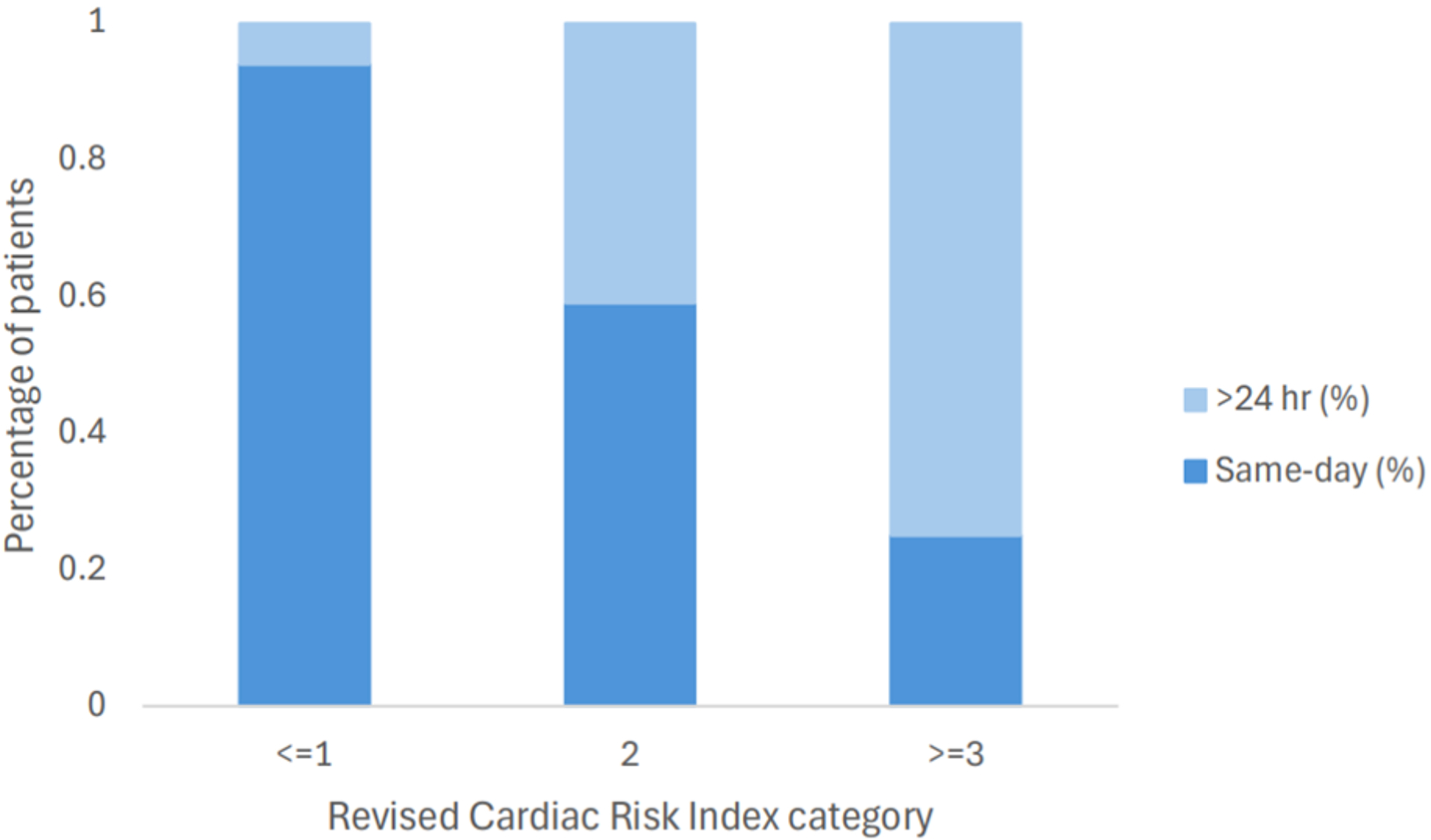
Distribution of discharge timing following major non-cardiac surgery by Revised Cardiac Risk Index (RCRI) category. Stacked bars represent the proportions of patients discharged on the same day and >24 hours after surgery.

**Table 4.**
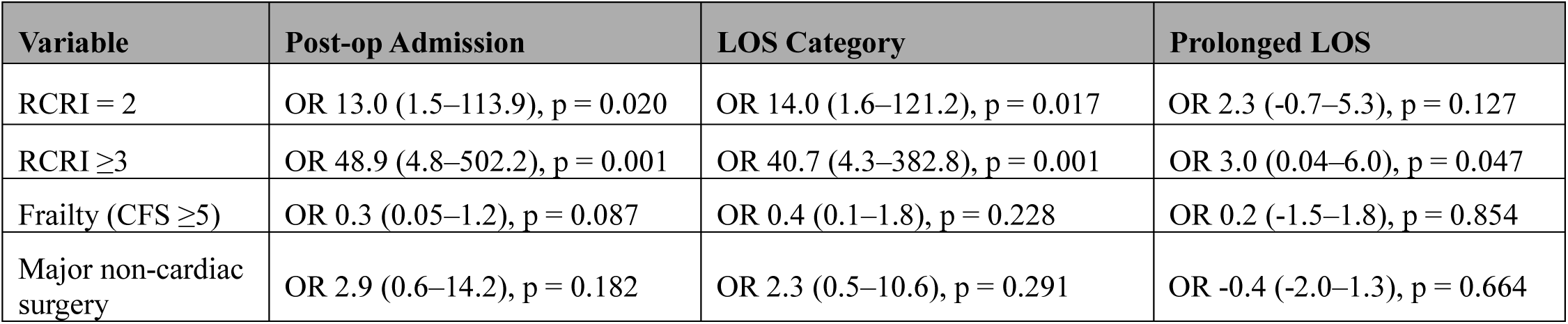
Association of RCRI, frailty, and major non-cardiac surgery with post-operative outcomes. Multivariable logistic regression was used for post-operative admission (≥24 hours), and ordinal logistic regression was used for length-of-stay (LOS) categories (same-day, 24–48 hours, >48 hours). Firth penalized logistic regression was applied for prolonged LOS (>48 hours) due to the small number of events. All models were adjusted for RCRI, frailty (CFS ≥5), and major non-cardiac surgery. CFS-clinical frailty scale, CI-confidence interval, OR-odds ratio, RCRI-revised cardiac risk index.

Length of stay showed a graded relationship with RCRI score. Among major non-cardiac procedures, same-day discharge occurred in 32 procedures (63%), 24–48 hours in 13 (25%), and >48 hours in six (12%). Ordinal logistic regression demonstrated that higher RCRI score was significantly associated with increasing LOS category: compared with RCRI score 0–1, RCRI score 2 had an adjusted OR of 14.0 (95% CI 1.6–121.1, p = 0.017), and RCRI score ≥3 had an adjusted OR of 40.7 (95% CI 4.3–382.9, p = 0.001), after adjusting for frailty and major non-cardiac procedures (**Figure 2**).

**Figure 2.**
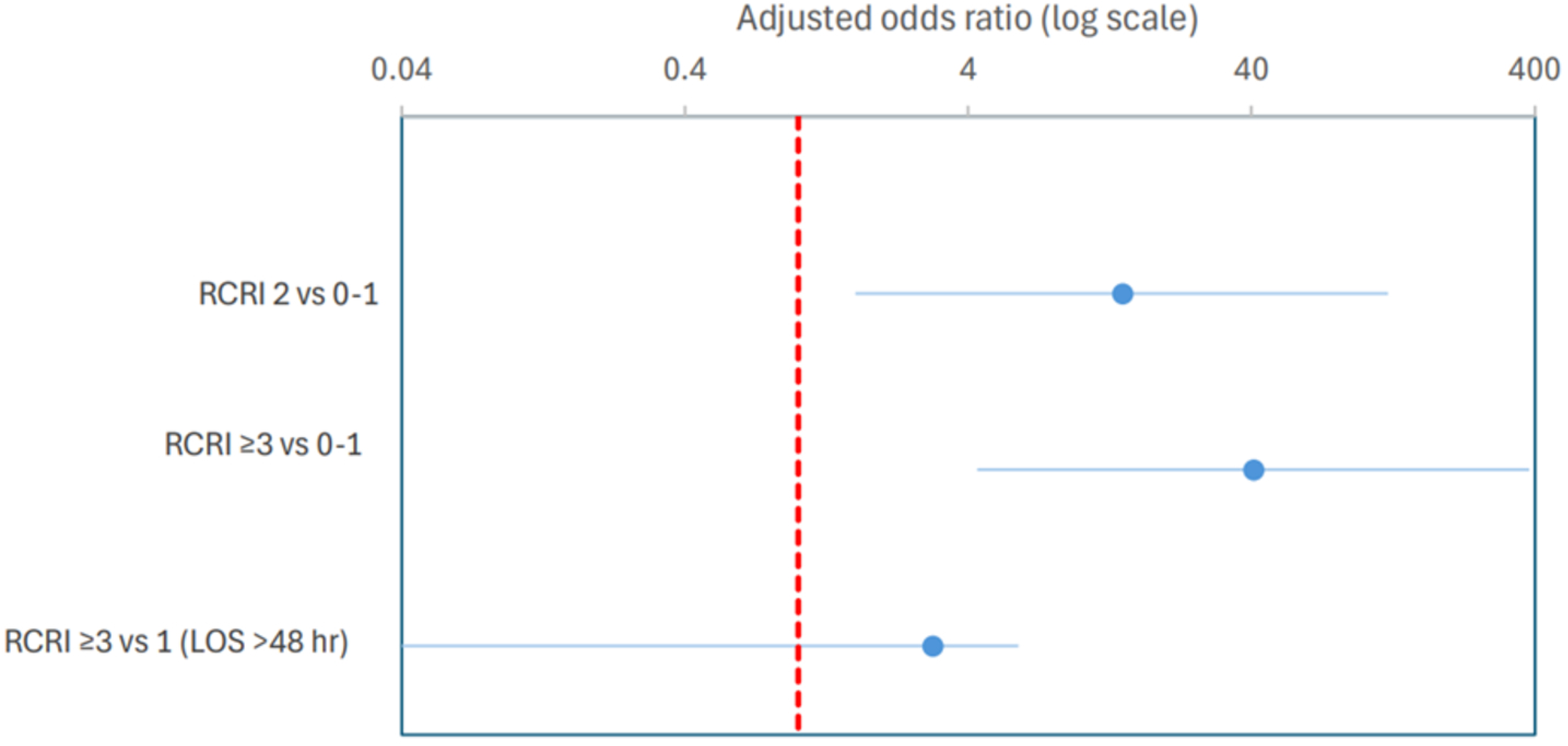
Forest plot of adjusted odds ratios (ORs) and 95% confidence intervals (CIs) for factors significantly associated with length of stay (LOS). Estimates were derived from ordinal logistic regression for LOS category analyses and from Firth penalized logistic regression for prolonged LOS (>48 hours). Only statistically significant adjusted associations are shown.

Prolonged hospitalization (LOS >48 hours) occurred in six patients. Using Firth penalized logistic regression, RCRI score ≥3 was significantly associated with prolonged hospitalization (adjusted OR 3.0, 95% CI 0.04–6.0, p = 0.047), while RCRI score 2 showed a non-significant trend. Sensitivity analysis using dichotomized RCRI score (≥3 vs <3) confirmed the robustness of the association. Frailty was common but was not associated with prolonged hospitalization. Adjustment for frailty and major non-cardiac surgery did not significantly change the estimates.

## DISCUSSION

The primary finding of this study is that the RCRI score retained utility for peri-operative risk stratification in a real-world ATTR-CM cohort undergoing major non-cardiac surgery. Higher RCRI scores, particularly RCRI ≥3, were associated with an increased requirement for postoperative hospital admission, longer postoperative LOS, and prolonged hospitalization beyond 48 hours, demonstrating a graded relationship across outcomes. The absolute number of MACE events and postoperative complications (according to the Clavien-Dindo classification system) was low overall, these findings occurred within an older and highly comorbid cohort generally considered at elevated peri-operative risk. Frailty was common in this real-world ATTR-CM population, although it was not independently associated with postoperative admission or prolonged hospitalization following adjustment. These findings are increasingly relevant given rising rates of ATTR-CM diagnosis and improving survival following the introduction of disease-modifying therapies, resulting in a growing number of ATTR-CM patients potentially presenting for surgical procedures.^4,5^

Peri-operative risk assessment in ATTR-CM remains inherently challenging due to the complex and multisystem nature of the disease. ATTR-CM is characterized by myocardial amyloid infiltration producing a restrictive physiology with impaired ventricular compliance, elevated filling pressures, and dependence on heart rate to maintain cardiac output in the context of a relatively fixed stroke volume.^3,18^ These hemodynamic abnormalities may render patients particularly vulnerable to peri-operative fluid shifts, vasodilation, and myocardial depression associated with anesthesia and surgery.^18^ In addition, impaired autonomic function may contribute to hypotension and hemodynamic instability, while ventricular stiffness and abnormal vascular coupling further limit cardiovascular reserve.^18^ Consequently, maintaining peri-operative hemodynamic stability in ATTR-CM patients may be especially difficult, particularly during major surgical procedures.

ATTR-CM patients also frequently exhibit various cardiovascular manifestations that may further increase peri-operative vulnerability. Atrial arrhythmias, conduction system disease, autonomic dysfunction, and aortic stenosis, are highly prevalent in this population and may independently contribute to peri-operative instability.^1,3,18,19^ The high prevalence of atrial fibrillation and long-term anticoagulation may also increase bleeding risk or stroke risk when interrupted for surgery. Despite these important disease-specific considerations, there remains a lack of robust peri-operative outcomes data in ATTR-CM, and no dedicated peri-operative risk stratification tools currently exist for this population. A prior peri-operative review of cardiac amyloidosis has similarly emphasized that conventional surgical risk models may incompletely capture the unique mechanisms contributing to peri-operative risk in ATTR-CM.^18^ This has also been highlighted in individual peri-operative case reports, including by Fernandes et al., who described the anesthetic and hemodynamic challenges encountered in an elderly ATTR-CM patient undergoing surgery following traumatic cervical spine injury.^19^ In this context, our findings suggest that while the RCRI score retains value in ATTR-CM patients undergoing surgery, important amyloid-specific determinants of peri-operative vulnerability likely remain unmeasured.

Despite the theoretical peri-operative vulnerability associated with ATTR-CM, the overall incidence of major postoperative complications was low overall. This may reflect careful patient selection, better understanding of ATTR-CM and its clinical sequelae or improved overall disease stability in an era of targeted disease-modifying therapy. Nevertheless, higher RCRI scores demonstrated a consistent graded association with postoperative admission and prolonged hospitalization, suggesting that the RCRI continues to provide informative risk stratification even within this high-risk population. Importantly, the association between higher RCRI scores and hospital stays exceeding 48 hours may be particularly clinically relevant, as this threshold likely reflects increased postoperative complexity requiring prolonged monitoring and greater peri-operative resource utilization. This is supported by peri-operative guideline-recommended postoperative surveillance pathways, which typically involve structured cardiac monitoring and biomarker assessment during the first 48-72 hours following surgery in high-risk patients identified by tools such as the RCRI.^7,8^ Accordingly, hospitalization beyond this period may be more indicative of unplanned postoperative complexity rather than routine monitoring.

These findings are broadly consistent with literature demonstrating that increasing RCRI scores are associated not only with cardiac complications, but also with longer hospitalization and greater postoperative morbidity across non-cardiac surgical populations.^11,20^ Similarly, studies in heart failure and cardiac surgery populations have identified frailty as an important predictor of prolonged hospitalization, postoperative complications, and mortality.^21,22^ Although frailty was prevalent in our cohort, it was not independently associated with postoperative outcomes following multivariable adjustment. This may relate to our smaller sample size, or suggest that conventional frailty measures such as the CFS may not fully capture peri-operative risk in ATTR-CM, and that cardiac-specific risk stratification tools such as the RCRI may provide complementary prognostic information in this population. These findings also highlight the potential value of developing a disease-specific peri-operative risk model in ATTR-CM that incorporate its unique pathophysiology.

### Limitations

This study has a single-centre, retrospective observational design; therefore, the presence of bias cannot be excluded. The limited sample size and low number of outcome/events reduced statistical power and may have prevented more detailed subgroup analyses. This was particularly relevant for rare outcomes, requiring the use of Firth penalized regression to address data sparsity. Additionally, while the RCRI was associated with postoperative outcomes in this cohort, it does not incorporate amyloid-specific disease features and may not fully capture all determinants of peri-operative risk in ATTR-CM.

## CONCLUSION

In this real-world cohort of ATTR-CM patients undergoing major non-cardiac surgery, while the overall risk of adverse outcomes was low, the RCRI demonstrated an association with postoperative admission and length of stay, including prolonged hospitalization beyond 48 hours. Given the complex multisystem nature of ATTR-CM, patients with higher RCRI scores may experience a more resource-intensive postoperative course. These findings suggest that the RCRI retains utility in ATTR-CM and may provide useful peri-operative risk stratification, while also highlighting the need for disease-specific risk models in this high-risk patient population.

## Acknowledgements

The authors acknowledge the nurses and staff of the Cardiac Amyloidosis Clinic and Amyloidosis Program of Calgary.

## Data Availability

Data from this study are available upon reasonable request to the corresponding author.

## Ethics Statement

This study was approved by the University of Calgary Research Ethics Board with a waiver of the requirement for patient informed consent.

### Patient Consent

The authors confirm that patient consent is not applicable to this study as many subjects were deceased or severely ill. The research ethics board waived the need for informed patient consent.

